# WATCH-SS: Developing a Trustworthy and Explainable Modular Framework for Detecting Cognitive Impairment from Spontaneous Speech*

**DOI:** 10.1101/2025.08.06.25333047

**Authors:** Sydney Pugh, Matthew Hill, Sy Hwang, Rachel Wu, Kuk Jang, Stacy Iannone, Karen O’Connor, Kyra O’Brien, Eric Eaton, Kevin Johnson

**Affiliations:** Department of Biostatistics, Epidemiology, and Informatics, Perelman School of Medicine, University of Pennsylvania, Philadelphia, PA USA; Department of Neurology, Perelman School of Medicine, University of Pennsylvania, Philadelphia, Pennsylvania, USA; Department of Computer and Information Science, School of Engineering and Applied Science, University of Pennsylvania, Philadelphia, PA USA; Department of Computer Engineering, Hongik University, Seoul, South Korea

**Keywords:** cognitive impairment, Alzheimer’s disease, dementia, natural language processing, large language model, machine learning

## Abstract

Early detection of cognitive impairment (CI) is critical for timely intervention in Alzheimer’s disease and AD-related dementias. To address this, we propose the Warning Assessment and Alerting Tool for Cognitive Health from Spontaneous Speech (WATCH-SS), a modular and explainable three-stage framework for detecting CI from a patient’s speech sample. The framework uses detectors for five linguistic and acoustic indicators of CI, aggregates their outputs into a set of clinically interpretable summary features, and uses a predictive model for CI classification. We consider multiple approaches to implementing these detectors that range from simple, computationally efficient methods suitable for real-time analysis to strong, resource-intensive methods, better for high accuracy offine analysis. On the DementiaBank ADReSS dataset, WATCH-SS achieved strong predictive performance (AUC = 80% on the test set). This work demonstrates that a modular, feature-based approach can achieve strong performance while providing a transparent diagnostic profile, representing a significant step towards a trustworthy and clinically-usable screening tool for primary care.

## 1. Introduction

As the global population ages, Alzheimer’s disease (AD) and AD-related dementias (ADRD) are becoming more prevalent.^1^ Yet, more than 50% of persons with AD/ADRD are undiagnosed,^2^ delaying crucial interventions that could improve quality of life. The first step in diagnosing AD/ADRD is detecting any degree of cognitive impairment (CI). Ideally, early detection should occur in primary care, where most patients first exhibit subtle signs of CI. Language production is highly sensitive to the neurodegenerative changes characteristic of AD/ADRD. Deficits in core cognitive domains—including semantic memory, executive function, and processing speed—manifest directly in a person’s speech patterns.^3–5^ Several of these signs manifest in conversations between patients and primary care providers (PCPs) during clinical visits—for example, when a patient repeats themselves several times during the visit or takes a long time to respond to the PCP’s questions. However, PCPs often struggle to recognize these clues in real time due to competing priorities, time constraints, and limited expertise.^6^

Speech data can be collected easily and non-invasively, making it an ideal medium for developing accessible tools for early-stage screening of CI. Recent advances in machine learning and natural language processing (NLP) have enabled the development of automated systems for detecting possible CI from speech.^7–9^ However, many state-of-the-art approaches rely on “black-box” deep learning models.^10–12^ While these systems can achieve high predictive accuracy, their opaque nature makes it impossible to understand the clinical reasoning behind a prediction, hindering trust and adoption in medical settings.

To address this gap, we propose the Warning Assessment and Alerting Tool for Cognitive Health for Spontaneous Speech (WATCH-SS), a modular framework designed to be a trustworthy, interpretable, and extensible tool for detecting CI due to suspected AD/ADRD from a patient’s speech sample. The framework leverages several detectors to identify indicators of CI within a speech sample and then aggregates the outputs into a clinically interpretable CI prediction. We demonstrate that our framework achieves strong predictive performance while providing a transparent diagnostic profile that explains the reasoning behind its predictions.

## 2. Related Work

Spontaneous speech, reliant on cognitive functions like attention and memory, is increasingly used as a non-invasive, cost-effective biomarker for detecting cognitive impairments. Clinically, speech is typically analyzed via structured speech tasks like the *Cookie Theft Picture Description* task from the Boston Diagnostic Aphasia Examination,^13^ verbal fluency tasks such as the Semantic Fluency Task (SFT) and Phonemic Fluency Task (PFT),^14^ and cognitive tests including the Mini-Mental State Examination (MMSE),^15^ the Montreal Cognitive Assessment (MoCA),^16^ and the Saint Louis University Mental Status (SLUMS) examination.^17^ While these assessments have been rigorously validated, they typically require in-person administration and expert interpretation, making them time-consuming and costly, which can delay diagnosis and treatment of neurodegenerative conditions like AD/ADRD.

Advances in machine learning offer the potential to overcome these limitations by enabling low-cost, remote, and early detection of cognitive decline through speech. They have revealed subtle linguistic and acoustic markers, expanded clinically relevant feature extraction, and enabled scalable, automated speech analysis.

To address these limitations, machine learning has been widely applied to automate the analysis of speech for detecting CI. Early works focused on training traditional models like Support Vector Machines (SVMs) on hand-crafted acoustic features (e.g., pause duration, speech rate)^18^ and lexical features (e.g., topic modeling).^19^ We refer the reader to Yang et al. ^20^ for an extensive survey of deep learning-based techniques. More recently, deep learning approaches using models like Convolutional Neural Networks (CNNs) and Long Short-Term Memory networks (LSTMs) have been used to learn patterns directly from audio.^21–23^ Transfer learning with large pre-trained models such as BERT, GPT, and wav2vec has also become a common strategy for extracting powerful text and audio embeddings for training various ML learning classifiers.^21,24–26^ These methods, often combined in multimodal or ensemble systems,^27,28^ have demonstrated high predictive accuracy but low explainability.

While other approaches have also explored training classifiers on interpretable speech features,^8,13,29,30^ they often use a mix of clinically interpretable and non-interpretable features. For example, these systems often incorporate acoustic features like GeMAPS or ComParE which are not easily parsed by clinicians. In contrast, WATCH-SS is designed to provide greater interpretability by focusing on high-level, clinician-interpretable features. This approach, coupled with external validation on spontaneous clinical speech, differentiates our work.

## 3. Methods

### 3.1. WATCH-SS

We propose WATCH-SS, a three-stage framework for detecting cognitive impairment from a patient’s speech sample (see Figure 1). The key advantages of our framework are its interpretability and modularity. Unlike “black-box” models, WATCH-SS provides a transparent patient diagnostic profile that breaks down the final CI prediction into the contributions of various indicators of CI, enabling a clinician to understand which specific cognitive deficits are driving the classification. Moreover, the modular architecture of WATCH-SS allows for easy extension or replacement of individual detectors.

**Fig. 1:**
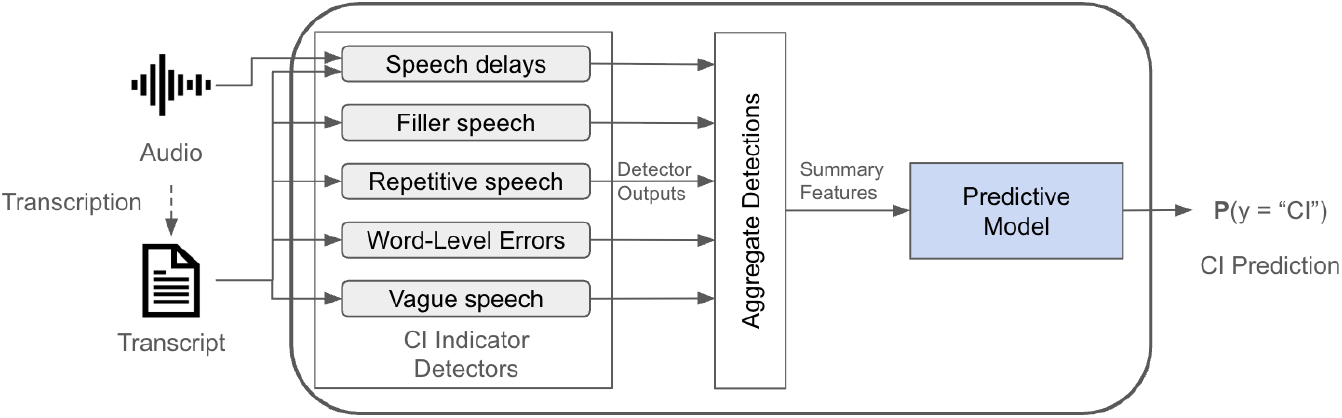
The WATCH-SS framework is a three-stage pipeline that (1) detects cognitive impairment (CI) indicators from audio and transcripts, (2) aggregates these detections into summary features, and (3) applies a predictive model to the features to generate a final prediction.

WATCH-SS receives as input an audio waveform and the corresponding diarized transcript of a speech sample. First, WATCH-SS runs multiple detectors in parallel to identify five key indicators of CI: filler speech, repetitive speech, substitution errors, vague speech, and speech delays. In the second stage, the system aggregates the detections into a set of clinically interpretable summary features and feeds them to a model to predict CI in the third stage.

WATCH-SS was implemented in Python (version 3.12.3) using Microsoft Azure Databricks. The code is available at https://github.com/kbjohnson-penn/WATCH-SS.

The remainder of this section describes the data used to develop WATCH-SS (Section 3.2), the detectors for indicators of CI (Section 3.3), the summary features extracted from these detections (Section 3.4), and finally, the predictive model for CI (Section 3.5).

### 3.2 Data

We developed and validated WATCH-SS using two datasets: the ADReSS dataset for detector and predictive model development, and the OBSERVER dataset for external validation.

#### 3.2.1. DementiaBank ADReSS Challenge Dataset

The ADReSS dataset^31,32^ consists of audio recordings and diarized transcripts of subjects with and without an AD diagnosis describing the Cookie Theft picture from the Boston Diagnostic Aphasia Exam.^33^ Transcripts were annotated using the CHAT coding system, a standardized protocol for transcribing conversational interactions and annotating its linguistic features.^34^ The training data consists of 108 subjects, while the test data consists of 48 subjects. Each dataset is balanced in age and gender and half of their subjects have an AD diagnosis.

We further partitioned the training data into training and development (dev) datasets using a 70/30 percent split balanced for age, gender, and number of AD subjects. Specifically, we first generated 100 candidate splits stratified by gender and AD label using different random seeds. Then, we selected the split that minimized the absolute difference in mean age across the resulting training and development sets. Table1 summarizes the resulting datasets.

**Table 1:**
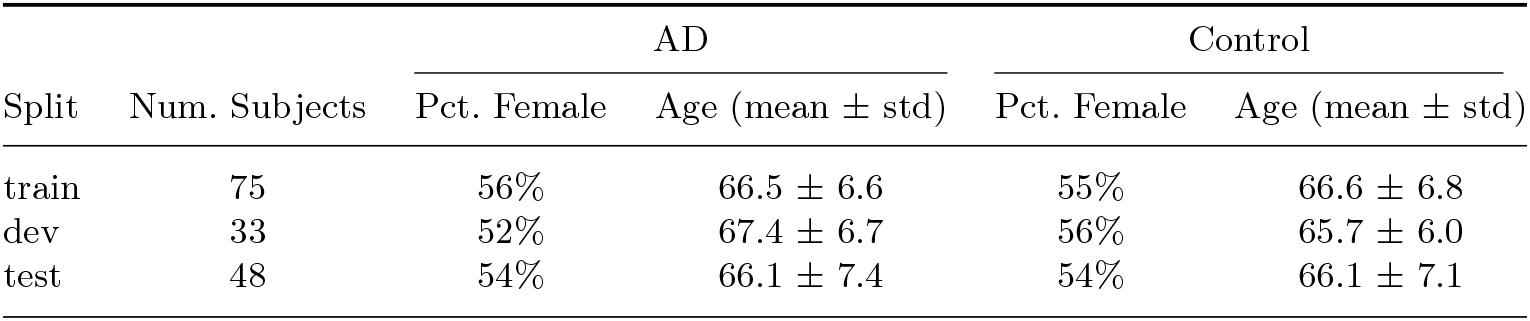
Characteristics of the ADReSS train, development (dev), and test datasets.

| Split | Num. Subjects | AD |  | Control |  |
| --- | --- | --- | --- | --- | --- |
| | | Pct. Female | Age (mean ± std) | Pct. Female | Age (mean ± std) |
| train | 75 | 56% | 66.5 ± 6.6 | 55% | 66.6 ± 6.8 |
| dev | 33 | 52% | 67.4 ± 6.7 | 56% | 65.7 ± 6.0 |
| test | 48 | 54% | 66.1 ± 7.4 | 54% | 66.1 ± 7.1 |

##### Data Pre-processing

We processed the CHAT transcripts to produce clean inputs to our detectors by: (1) converting pause annotations to a “[silence]” token, (2) converting event annotations to “[*<*event *>*]” tokens, (3) converting inaudible annotations to “[inaudible]” tokens, and (4) removing all other CHAT annotations. Then, we derived ground-truth labels from the original CHAT annotations for all patient utterances. An utterance was labeled positive for a given indicator if it met the following criteria:

- **Filler speech:** The utterance contained any word marked with the filler prefix (“&”).
- **Repetitive speech:** The utterance contained the repetition annotation (“[/]”).
- **Substitution errors:** The utterance contained the word level error code (“[*]”), including all its subtypes (*e*.*g*., “[* s]” for semantic paraphasias or “[* n]” for neologisms).
- **Vague speech:** The utterance contained the annotation for empty speech (“[+ es]”) or circumlocution (“[+ cir]”).
- **Speech delays:** The utterance contained a pause annotation (“(.)”, “(..)”, or “(…)”).

The prevalence of these labels across the data splits is summarized in Table 2.

**Table 2:**
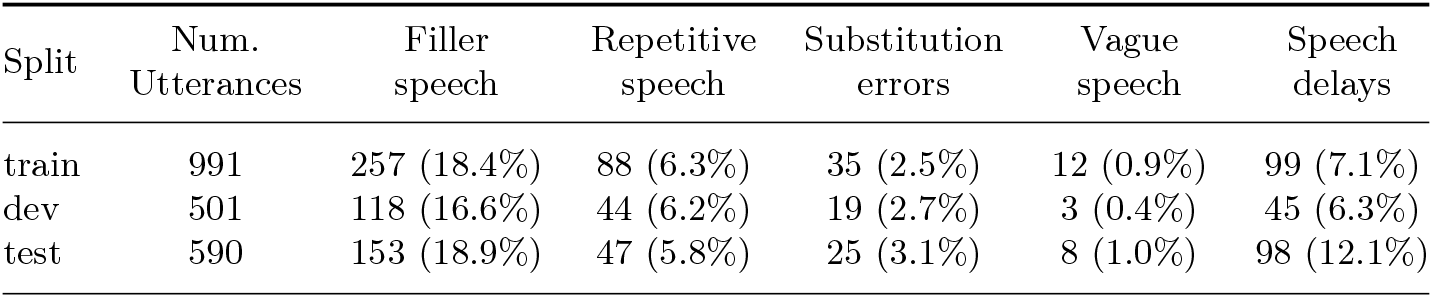
Prevalence of cognitive impairment (CI) indicators in the ADReSS train, development (dev), and test datasets. Values are the counts and proportions of positively-labeled utterances.

#### 3.2.2. Penn OBSERVER Repository Dataset

The OBSERVER Repository was developed by the University of Pennsylvania to capture and support detailed, multimodal analyses of real outpatient clinical visits.^35^ For this work, we used the visit recordings, diarized manual transcripts, and corresponding Electronic Health Record (EHR) data from the repository. We preprocess the audio from the recordings to reduce any background noise using Adobe Premiere Pro’s audio enhancement tools. ^a^

To construct our external validation dataset, we first selected the earliest recorded visit for all patients aged 50 or older (74 patients in total). Next, to identify patients with CI we reviewed each patient’s EHR data available up to the time of visit for mention of “cognitive impairment”, “Alzheimer’s”, or “dementia” in the diagnosis list, problem list, medical history, or encounter notes. This process identified 14 CI patients, two of which we excluded because their CI was not due to suspected AD/ADRD. To create a balanced case-control dataset, we downsampled the cognitively normal (“CN”) pool to 12 patients that best balances age and gender with the CI group. Table 3 summarizes the dataset.

**Table 3:**
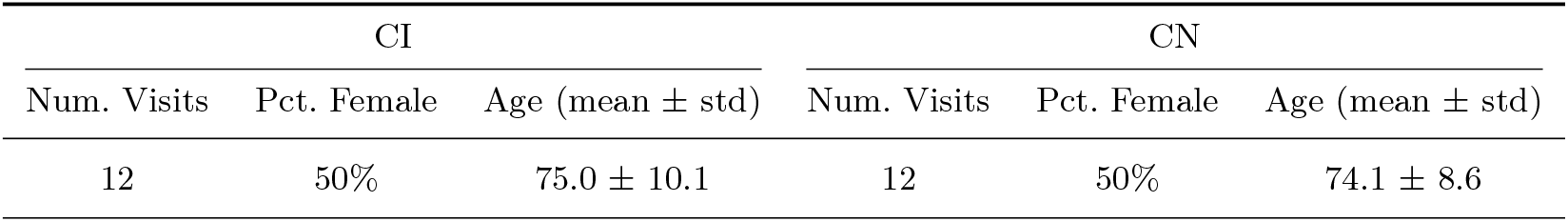
Characteristics of the OBSERVER Repository dataset.

| CI |  |  | CN |  |  |
| --- | --- | --- | --- | --- | --- |
| Num. Visits | Pct. Female | Age (mean ± std) | Num. Visits | Pct. Female | Age (mean ± std) |
| 12 | 50% | 75.0 ± 10.1 | 12 | 50% | 74.1 ± 8.6 |

### 3.3. Detectors for Indicators of Cognitive Impairment

We developed detectors for five key indicators of CI. Four are linguistic indicators extracted from the transcript–filler speech, repetitive speech, substitution errors, and vague speech– whereas the fifth indicator, speech delays, is an acoustic indicator derived from the audio.

A significant challenge to developing specialized detectors for these CI indicators is the lack of large, publicly available annotated datasets. To overcome this, we focus on low-cost approaches that are straightforward to implement and do not require extensive amounts of annotated data for task-specific fine-tuning. We implemented a single detector for speech delays, as their identification relies on silence detection–a well-established audio signal processing task. For the linguistic indicators, however, we implement and compare two primary approaches:

1. **Traditional NLP**: Computationally efficient methods for capturing well-defined linguistic patterns that serve as strong, high-recall baselines.
2. **LLM Prompting**: To evaluate the “out-of-the-box” capability of modern, pre-trained LLMs for detecting linguistic indicators of CI we use zero- and few-shot prompting.

To implement the LLM-based detectors, we created a general-purpose prompt template, shown in Figure 2, that we adapt for each specific CI indicator. The prompt instructs the LLM to adopt the persona of a neurologist, provides a detailed definition of the target CI indicator with rules for detection, and enforces a JSON output format. We iteratively refined all prompts for linguistic indicators to maximize the F1-score on the ADReSS training dataset. For the fewshot prompts, examples were selected from the ADReSS development dataset based on an error analysis of the zero-shot version of the prompt. All detectors use OpenAI’s GPT-4o LLM ^b^, which has shown promising performance on clinical tasks like speech analysis.^36–38^ To ensure reproducibility and reduce non-determinism, we fix the LLM’s *temperature* to 0 and *top p* to 1. These approaches offer different balances between detection accuracy and computational efficiency, allowing WATCH-SS to be configured with either fast, lightweight detectors for realtime screening or more computationally intensive, high-accuracy detectors for offine analysis.

**Fig. 2:**
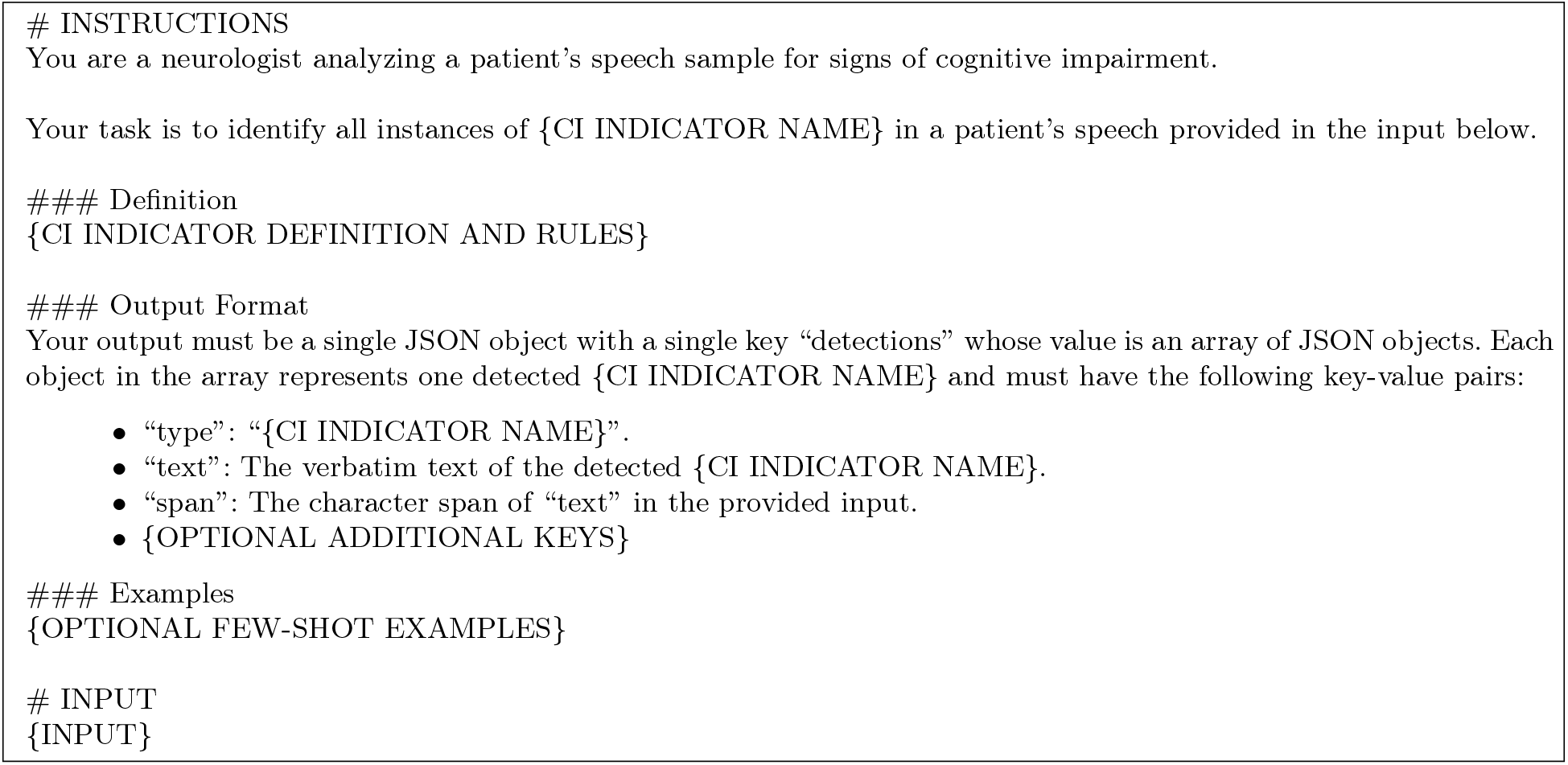
LLM-based detector prompt template.

The following subsections detail the clinical motivation for each indicator and the specific implementation of these detector types.

#### 3.3.1. Filler Speech

Fillers are sounds, words, or phrases used to improve speech planning, often as an alternative to silent pauses. Frequent use of fillers is a well-documented linguistic indicator of word-finding difficulties and increased cognitive load associated with CI and AD.^5^

##### NLP Baseline: Keyword Search Detector

Our baseline for filler speech is a simple, case-insensitive keyword search that processes each utterance and returns the text and character span for any matches. We use the spaCy Python library (version 3.8.7),^39^ with the pre-trained English model pipeline optimized for CPU “en core web md”, for tokenization and keyword matching. To determine the optimal filler keyword set, we evaluated combinations of the keyword categories defined in Table A1 on the ADReSS training data. The best-performing set was a combination of the “Sounds” and “Uncommon Letters” sets, which achieved the highest F1-score against filler speech labels (see Appendix A for the full results).

##### LLM-Based Detector

We use a zero-shot prompting strategy to implement the LLM-based detector for fillers. The detector processes the transcript utterance-by-utterance, as the context needed to identify a filler is typically contained within a single utterance. The prompt, provided in Appendix B, gives the LLM a simple description of fillers and sets explicit rules to avoid detecting event tags and repetitions. Due to the high performance achieved with the zero-shot approach, few shot examples were not added to the prompt.

#### 3.3.2. Repetitive Speech

Word repetition is a common speech pattern for individuals with CI and dementia.^40^ This pattern can be a manifestation of several underlying cognitive issues, including stuttering, word-finding difficulty, perseveration (the inability to switch from a completed thought), or an individual may not remember what they previously said in a conversation.^5^

##### NLP Baseline: Unigram Analysis

Our baseline detector for word repetition is based on a unigram analysis. The detector processes each utterance from a transcript by tokenizing it and then iterates through the tokens, checking for matches against the previous *K* seen tokens (unigrams). For each match, a JSON object with four key-value pairs: the”type” of detection (*i*.*e*., “repetition”), the “text” that is repeated, and the character span of each occurrence of the repeated text within the utterance, “span” and “span2”. As before, we use the spaCy library for tokenization. The detector has two key hyperparameters: the window size (*K*), which controls the distance for a match, and the comparator function, which determines the type of repetition being detected (*e*.*g*., verbatim repetitions or deeper semantic repetitions).

We optimized these hyperparameters to maximize the F1-score against the repetition labels on the ADReSS training dataset, and found a window size of two tokens with an exact match comparator function to be the best performing (see Appendix C for the full results).

##### LLM-Based Detector

We started with the zero-shot prompting strategy for the word repetitions LLM-based detector with utterance-by-utterance processing. The prompt (provided in Appendix D) described repetition as involuntary, verbatim repeats of whole words, and includes an additional key for the JSON output format, “span2”, to capture the span of the second occurrence of the word. To improve the low precision of this high-recall zero-shot prompt, we added few-shot examples from the ADReSS development data that specifically target common failure cases, such as flagging distant repetitions and revisions of word forms.

#### 3.3.3. Substitution Errors

Substitution errors are a phenomenon where an individual replaces an intended word with an unintended word. While occasional errors can occur in cognitively normal individuals, frequent and consistent errors can indicate CI, and in more severe cases, aphasia–a language disorder resulting from damage to the brain.^5,41–44^ These errors manifest in several ways, including phonemic, semantic, and neologistic paraphasias, morphological errors, and intra-word dysfluencies.

##### NLP Baseline: MLM-Based Detector

We leverage the BERT Masked Language Model (MLM)^45^ to identify substitution errors based on the contextual predictability of each word spoken by the patient. Since these errors are contextually inappropriate words, we hypothesize that a MLM will assign them a very low probability, making them detectable as outliers.

Based on this hypothesis, the detector systematically quantifies the unpredictability of each word spoken by the patient in the transcript. For each word in the transcript, we mask it and then use the MLM to predict the masked word, which generates a probability distribution over its entire vocabulary. From this distribution, we compute two complementary metrics to assess the actual masked word’s contextual fit: *normalized entropy*, which measures the overall uncertainty of the model’s prediction (normalized by vocabulary size); and the *concentration ratio*, which measures how confident the model is in its top guesses. These scores are then combined into a single fusion score, and the word is flagged as a substitution error if this score exceeds a predetermined percentile threshold. We optimized this threshold on the ADReSS training dataset, finding that the 90th percentile maximized the F1-score against ground-truth substitution error labels (see Appendix E for the full results).

##### LLM-Based Detector

Our prompt for the substitution error LLM-based detector leverages the zero-shot prompting strategy. Similar to the baseline, this detector processes the entire transcript at once in order to leverage sufficient context for identifying substitution errors. The prompt (provided in Appendix F) includes detailed definitions and examples for the each of the aforementioned types of substitution errors. To improve the model’s reasoning, we also employ a Chain-of-Thought approach by adding a “justification” key to the JSON output, instructing the LLM to provide a brief explanation for each word it flags. We also tried adding few-shot examples from the ADReSS development based on an error analysis of the zero-shot prompt to improve performance, but this yielded minor gains in precision that were offset by a larger drop in recall.

#### 3.3.4. Vague Speech

Decreased speech content–that is, speech that is correct but conveys little to no meaning–is a powerful indicator of cognitive decline driven by overuse of vague language. As a person’s cognitive status declines, their knowledge base for concepts and word meanings deteriorates, and they struggle to retrieve specific words from memory. Consequently, they may overuse unspecific referents for more specific words or talk around words or concepts (*i*.*e*., circumlocution).^5^

##### NLP Baseline: Keyword Search Detector

We implement a keyword search to detect specific vague terms and phrases, using the same implementation as the filler keyword detector from Section 3.3.1. To determine the optimal keyword set, we experimented with the keyword categories defined in Table G1 using the ADReSS training dataset. The “Non-Specific Referents” keyword set achieved the maximum F1-score (see Appendix G for the full results).

##### LLM-Based Detector

Our LLM-based detector for vague speech uses a few-shot prompt. Error analysis of the baseline’s high recall but low precision performance suggested that the context in which terms are used is crucial for determining vagueness. Hence, this detector processes the entire transcript at once. The prompt (see Appendix H) provides the LLM with a detailed definition of vague speech and, crucially, includes few-shot examples that teach the model to ignore general terms when their meaning is clarified by the surrounding context or when they represent normal conversational patterns rather than word-finding difficulty.

#### 3.3.5. Speech Delays

Speech delays are unfilled pauses in speech that cannot be attributed to normal respiratory breaks. These delays are strongly associated with several cognitive deficits characteristic of AD, including memory deficits, slowed cognitive processing and impaired executive function.^3^

##### Silence Detector

To identify speech delays, silences in the audio were detected using the <monospace>detect_silence</monospace> function from the Pydub Python library (version 0.25.1),^46^ which identifies excerpts of audio quieter than the specified threshold. This function relies on two primary hyperparameters: the silence threshold, which sets the maximum decibel level relative to full scale (dBFS) for silence in audio, and the minimum silence length, which determines the minimum duration in milliseconds for an audio segment to be considered silence. The <monospace>detect_silence</monospace> function returns a list of tuples, each containing the start and end times (in milliseconds) of detected silent segments. We tuned the silence threshold on the ADReSS training data–selecting −55 dBFS for its high recall–while fixing the minimum silence length to 10ms to capture all potential pauses for subsequent feature extraction (see Appendix I for full details).

### 3.4. Summary Features

The detections for each of our five CI indicators are aggregated into a set of clinically interpretable summary features, which are defined in Table 4. To assess the individual utility of each feature, we performed a univariate analysis on the ADReSS training data (see Appendix J).

**Table 4:**
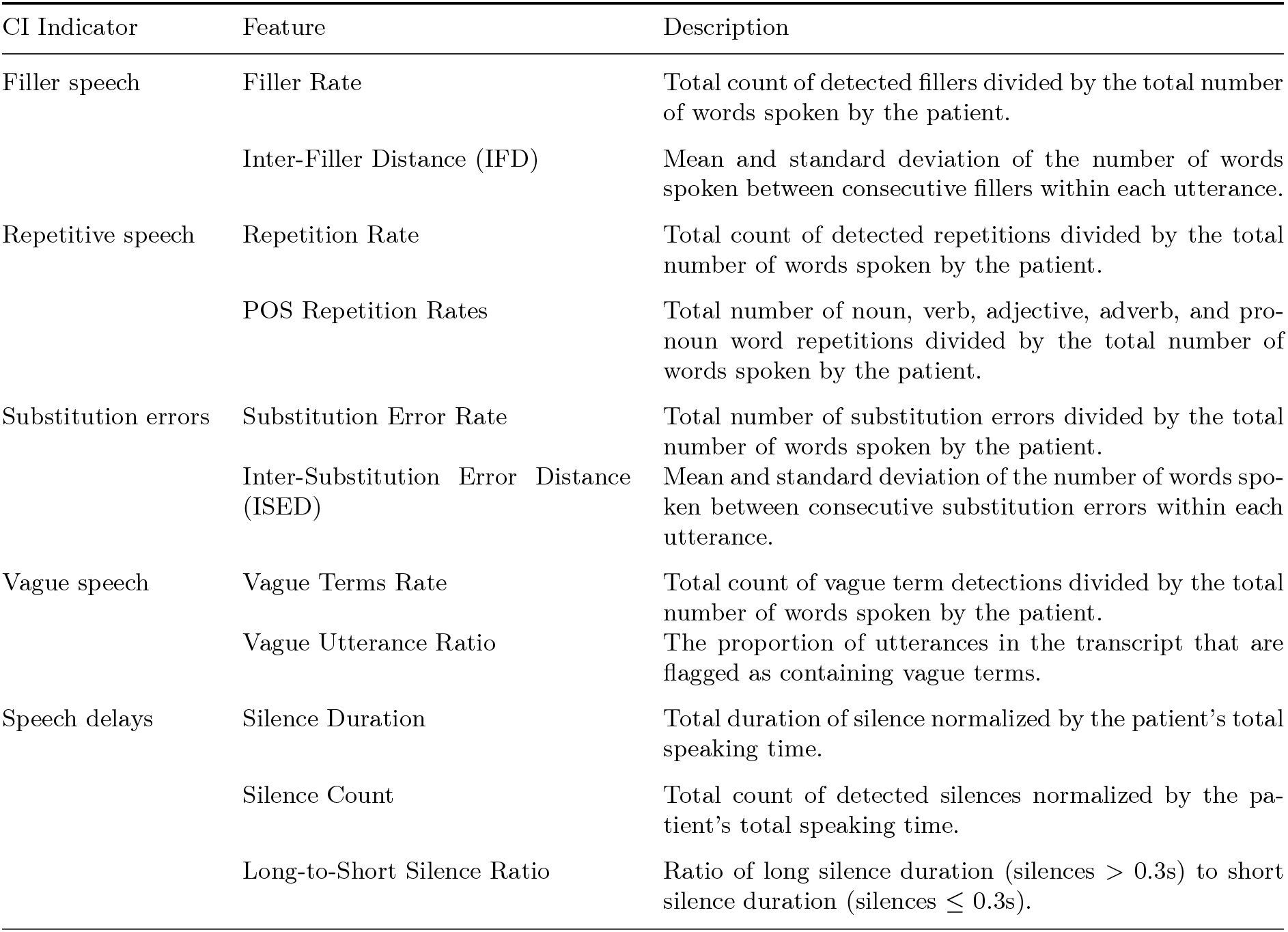
Summary of features for our five indicators of cognitive impairment (CI).

### 3.5. Predictive Model for Cognitive Impairment

To predict CI due to suspected AD/ADRD, we first computed the summary feature set for the ADReSS data using the outputs from our traditional NLP baseline detectors. These detectors were chosen for their practical advantages in a real clinical setting. On this feature set, we then experimented with a diverse set of machine learning models: logistic regression, random forest, and Histogram-based Gradient Boosting Classifier (HGBC), LightGBM, XGBoost, KNearest Neighbors (KNN), and support vector machine (SVM). Prior to training, we first dropped the determiner repetition rate feature because it had zero variance in the ADReSS data. Then, all remaining features were scaled to a [0, 1] range using a MinMaxScaler fitted only on the training data. While we also explored dimensionality reduction using t-distributed Stochastic Neighbor Embedding (t-SNE) and Principal Component Analysis (PCA), it did not consistently improve performance, so the full feature set was used for all models.

We used two cross-validation (CV) strategies: leave-one-out (LOO) and repeated stratified k-fold (RSKF) with 10 folds and 10 repeats. Cross-validation folds were created at the subject level to prevent data leakage between training and validation data. For each strategy, predictions on the validation sets across all training folds were ensembled using a hard voting approach to determine the final prediction. The hyperparameters for each model were tuned manually to optimize the average F1-score across CV strategies on the ADReSS training dataset. We implemented all model training using the scikit-learn Python library (version 1.4.2).^47^

After identifying the best configuration for each model type, we selected the overall best-performing model, LightGBM, based on its superior training CV performance. The model is configured with a binary objective, 10,000 estimators with early stopping patience of 50 rounds, maximum tree depth of 1, 12 data samples minimum per one leaf, L1 regularization, and restricting the model to use 20% of the features before training each tree. We focus on this LightGBM model in our main results and present the performance of all models in Appendix K.

## 4. Results

### 4.1. Performance of the Detectors for Cognitive Impairment Indicators

#### 4.1.1. Linguistic Indicators

Table 5 reports the performance of the NLP baseline and LLM-based detectors for the linguistic CI indicators on the ADReSS test data. The results show a clear distinction based on the complexity of the task. For indicators defined by specific lexical items like Filler Speech and Repetitive Speech, the NLP baselines achieved a better balance in precision and recall, while the LLM-based detectors consistently achieve high recall at the cost of lower precision. For example, the filler keyword search detector had a balanced and high precision and recall (94.1% and 93.5%, respectively), while the LLM’s tendency to over-predict resulted in a lower F1.

**Table 5:**
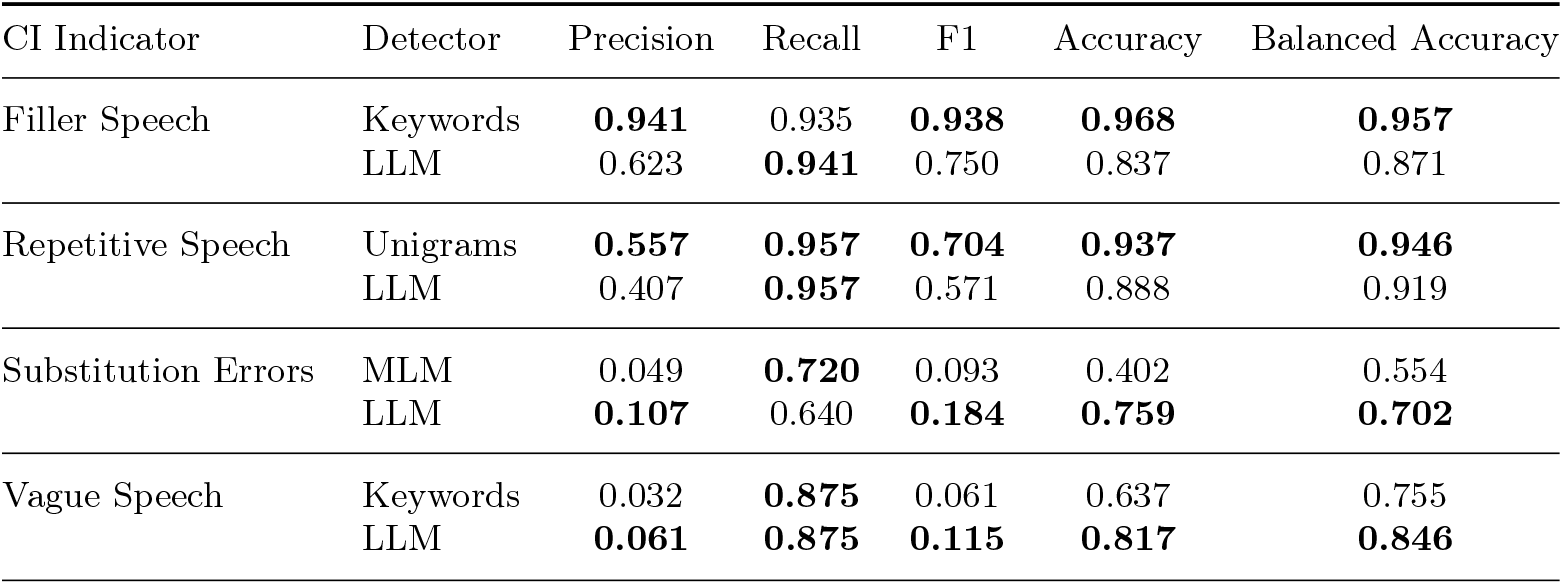
Performance comparison of natural language processing and LLM-based detectors for the linguistic indicators of cognitive impairment (CI) on the ADReSS test data. The best scores for each indicator are highlighted in boldface.

Conversely, for the more semantically complex tasks of substitution errors and vague speech, the LLM-based detectors demonstrated superior performance. For substitution errors, the LLM achieved a significantly higher F1-score (18.4%) than the MLM baseline (9.3%), driven by a notable improvement in precision. However, it is important to note that the low prevalence of positively-labeled utterances makes evaluation challenging and all of these detectors demonstrated very low precision.

#### 4.1.2. Speech Delays

The performance of the silence detector for identifying speech delays in the ADReSS test data is presented in Figure 3. The figure shows the trend in precision, recall, F1-score, and accuracy across a range of choices for the minimum silence length hyperparameter while, using the optimal −55 dBFS silence threshold that was determined on the ADReSS training data.

**Fig. 3:**
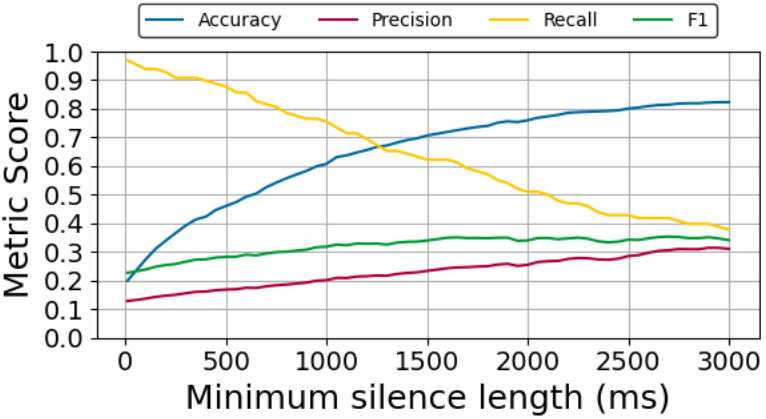
Performance of silence detector for identifying speech delays in the ADReSS test data.

As the minimum silence length increases, the detector becomes more conservative, only flagging long pauses, causing precision to steadily increase while the recall falls. The highest F1-score (35%) was achieved with a minimum silence length of 1,950ms.

### 4.2. Performance of the Cognitive Impairment Prediction Model

Figure 4 shows the performance characteristics of the best performing CI prediction model on the ADReSS training and test datasets. The ROC curves and AUC estimates demonstrate the model’s strong ability to discriminate between the cognitively impaired and normal groups. The model achieves a near-perfect AUC of 95% (CI: [0.905, 0.976]) on the training data, and generalized fairly well on the test data (AUC = 80%, CI: [0.682, 0.904]). The model also demonstrates strong F1-scores on both datasets, 87% (CI: [0.792, 0.935]) and 77% (CI: [0.647, 0.867]), on the training and test data, respectively. We anticipated a drop in performance due to a small degree of overfitting to the small training sample size. Nonetheless, the strong train and test performance demonstrates that a model is able to learn meaningful patterns for identifying CI, even from features derived from detectors that often favor high recall over precision.

**Fig. 4:**
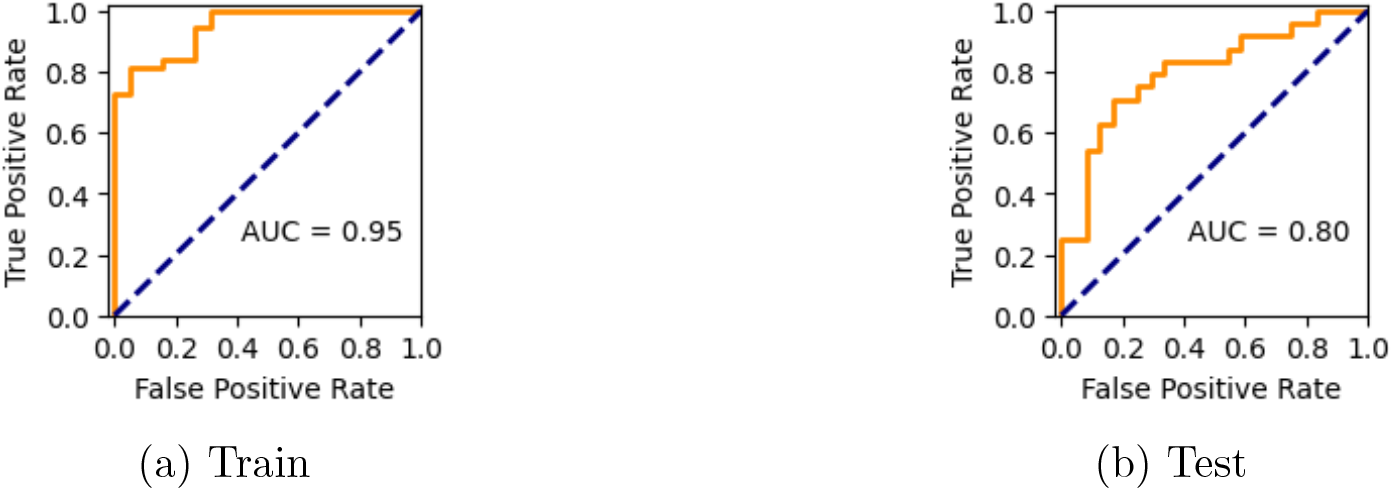
Performance characteristics of the CI prediction model on the ADReSS datasets.

### 4.3. External Validation of WATCH-SS

We performed an external validation of WATCH-SS on the OBSERVER dataset to assess its viability as an ambient CI screening tool for real-world primary care settings. The CI prediction model’s performance on this dataset was lower than on the ADReSS data, achieving an AUC of 63.2% (CI: [0.427, 0.829]) and F1-score of 31.6% (CI: [0.111, 0.526]).

## 5. Discussion

In this paper, we described and evaluated WATCH-SS, a modular framework designed for trustworthy and interpretable detection of cognitive impairment (CI) from a patient’s speech sample. Our results show that WATCH-SS can effectively discriminate between cognitively impaired and cognitively normal speech, achieving strong predictive performance on the ADReSS test set (AUC = 80%). A key finding is that this performance was achieved using features derived from the outputs of simple, computationally efficient NLP-based detectors. The success of the model, even with features from detectors that often favor high recall over high precision, highlights the viability of our framework as an ambient CI screening tool for primary care.

It is crucial, however, to interpret the performance of WATCH-SS in the context of several study and dataset limitations. A primary limitation is the small sample sizes, particularly for the external validation dataset. While the model performs well on the ADReSS dataset, its reduced performance on the OBSERVER dataset highlights the need for further validation on larger and more diverse clinical populations. Furthermore, the ground-truth CHAT annotations in the ADReSS dataset can be subjective and inconsistent. For instance, the CHAT manual lacks a strict, duration-based definition for speech delays, and the prevalence of annotated vague speech and substitution errors is extremely low. These limitations likely lead to an underestimation of our detectors’ true performance and highlight a broader challenge in the field. In principle, our detectors could be reused to address this by generating a large, weakly-annotated dataset of transcripts. Despite the limitations, the primary contribution of WATCH-SS lies in its modular and explainable framework, which aligns with clinician-interpretable indicators.

Our work highlights several avenues for future research. The modularity of WATCH-SS allows for the straightforward enhancements to our current detectors. For example, we plan to explore a generator-critic approach to improve the precision of our high-recall detectors. This involves using our current detectors as generators to produce candidate detections, which would then be filtered by an LLM “critic” to identify the most clinically relevant instances. However, the most critical next step is to further assess the framework’s real-world viability. This requires an evaluation of the framework using real-time Automatic Speech Recognition.^48–50^ Furthermore, our external validation on the OBSERVER dataset revealed that the shorter, more fragmented patient utterances in clinical visit dialogues are insufficient speech samples. We plan to test simple, open-ended prompts (e.g., “Describe your typical day”) that can easily be added to a clinical workflow to elicit better speech samples. These future steps are critical for making WATCH-SS a robust and trustworthy tool for clinical practice.

## Data Availability

The ADReSS dataset is available from the DementiaBank database at https://talkbank.org/dementia/ADReSS-2020/index.html. The OBSERVER dataset is available from the University of Pennsylvania at https://observer.med.upenn.edu/.

https://observer.med.upenn.edu/

## Supplementary Material

All appendices can be found at https://github.com/kbjohnson-penn/WATCH-SS/blob/main/supplementary_material.md.

## Footnotes

a https://www.adobe.com/products/premiere.html

b Experiments involving LLMs were conducted using HIPAA-compliant services available through the Microsoft Azure AI platform to ensure patient data privacy.

