## Supplementary Material for "WATCH-SS: Developing a Trustworthy and Explainable Modular Framework for Detecting Cognitive Impairment from Spontaneous Speech*"

Early detection of cognitive impairment (CI) is critical for timely intervention in Alzheimer's disease and AD-related dementias. To address this, we propose the Warning Assessment and Alerting Tool for Cognitive Health from Spontaneous Speech (WATCH-SS), a modular and explainable three-stage framework for detecting CI from a patient's speech sample. The framework uses detectors for five linguistic and acoustic indicators of CI, aggregates their outputs into a set of clinically interpretable summary features, and uses a predictive model for CI classification. We consider multiple approaches to implementing these detectors that range from simple, computationally efficient methods suitable for real-time analysis to strong, resource-intensive methods, better for high accuracy offline analysis. On the DementiaBank ADReSS dataset, WATCH-SS achieved strong predictive performance (AUC = 80% on the test set). This work demonstrates that a modular, feature-based approach can achieve strong performance while providing a transparent diagnostic profile, representing a significant step towards a trustworthy and clinically-usable screening tool for primary care.

Keywords: cognitive impairment, Alzheimer's disease, dementia, natural language processing, large language model, machine learning

---

*Research reported in this publication was supported by the National Institute On Aging of the National Institutes of Health under Award Number P30AG073105, and by the University of Pennsylvania ASSET Center under the Collaborative Research in Trustworthy AI for Medicine grant. The content is solely the responsibility of the authors and does not necessarily represent the official views of the National Institutes of Health.

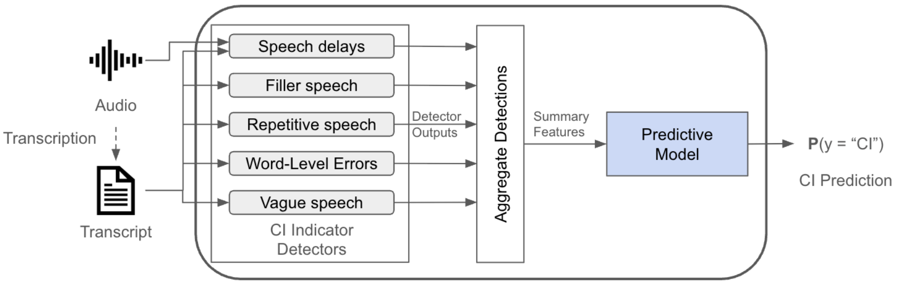


Fig. 1: The WATCH-SS framework is a three-stage pipeline that (1) detects cognitive impairment (CI) indicators from audio and transcripts, (2) aggregates these detections into summary features, and (3) applies a predictive model to the features to generate a final prediction.

The prevalence of these labels across the data splits is summarized in Table 2.

Table 1: Characteristics of the ADReSS train, development (dev), and test datasets.

| | | AD | | Control | |
|---|---|---|---|---|---|
| Split | Num. Subjects | Pct. Female | Age (mean ± std) | Pct. Female | Age (mean ± std) |
| train | 75 | 56% | 66.5 ± 6.6 | 55% | 66.6 ± 6.8 |
| dev | 33 | 52% | 67.4 ± 6.7 | 56% | 65.7 ± 6.0 |
| test | 48 | 54% | 66.1 ± 7.4 | 54% | 66.1 ± 7.1 |

Table 2: Prevalence of cognitive impairment (CI) indicators in the ADReSS train, development (dev), and test datasets. Values are the counts and proportions of positively-labeled utterances.

| Split | Num. Utterances | Filler speech | Repetitive speech | Substitution errors | Vague speech | Speech delays |
|---|---|---|---|---|---|---|
| train | 991 | 257 (18.4%) | 88 (6.3%) | 35 (2.5%) | 12 (0.9%) | 99 (7.1%) |
| dev | 501 | 118 (16.6%) | 44 (6.2%) | 19 (2.7%) | 3 (0.4%) | 45 (6.3%) |
| test | 590 | 153 (18.9%) | 47 (5.8%) | 25 (3.1%) | 8 (1.0%) | 98 (12.1%) |

Table 3: Characteristics of the OBSERVER Repository dataset.

| CI | | | CN | | |
|---|---|---|---|---|---|
| Num. Visits | Pct. Female | Age (mean ± std) | Num. Visits | Pct. Female | Age (mean ± std) |
| 12 | 50% | 75.0 ± 10.1 | 12 | 50% | 74.1 ± 8.6 |

###### 3.2.2. *Penn OBSERVER Repository Dataset*

[b]Experiments involving LLMs were conducted using HIPAA-compliant services available through the Microsoft Azure AI platform to ensure patient data privacy.

```
# INSTRUCTIONS
You are a neurologist analyzing a patient's speech sample for signs of cognitive impairment.

Your task is to identify all instances of {CI INDICATOR NAME} in a patient's speech provided in the input below.

### Definition
{CI INDICATOR DEFINITION AND RULES}

### Output Format
Your output must be a single JSON object with a single key "detections" whose value is an array of JSON objects. Each object in the array represents one detected {CI INDICATOR NAME} and must have the following key-value pairs:

  • "type": "{CI INDICATOR NAME}".
  • "text": The verbatim text of the detected {CI INDICATOR NAME}.
  • "span": The character span of "text" in the provided input.
  • {OPTIONAL ADDITIONAL KEYS}

### Examples
{OPTIONAL FEW-SHOT EXAMPLES}

# INPUT
{INPUT}
```

Fig. 2: LLM-based detector prompt template.

Table 4: Summary of features for our five indicators of cognitive impairment (CI).

| CI Indicator | Feature | Description |
|---|---|---|
| Filler speech | Filler Rate | Total count of detected fillers divided by the total number of words spoken by the patient. |
| | Inter-Filler Distance (IFD) | Mean and standard deviation of the number of words spoken between consecutive fillers within each utterance. |
| Repetitive speech | Repetition Rate | Total count of detected repetitions divided by the total number of words spoken by the patient. |
| | POS Repetition Rates | Total number of noun, verb, adjective, adverb, and pronoun word repetitions divided by the total number of words spoken by the patient. |
| Substitution errors | Substitution Error Rate | Total number of substitution errors divided by the total number of words spoken by the patient. |
| | Inter-Substitution Error Distance (ISED) | Mean and standard deviation of the number of words spoken between consecutive substitution errors within each utterance. |
| Vague speech | Vague Terms Rate | Total count of vague term detections divided by the total number of words spoken by the patient. |
| | Vague Utterance Ratio | The proportion of utterances in the transcript that are flagged as containing vague terms. |
| Speech delays | Silence Duration | Total duration of silence normalized by the patient's total speaking time. |
| | Silence Count | Total count of detected silences normalized by the patient's total speaking time. |
| | Long-to-Short Silence Ratio | Ratio of long silence duration (silences $>$ 0.3s) to short silence duration (silences $\leq$ 0.3s). |

Table 5: Performance comparison of natural language processing and LLM -based detectors for the linguistic indicators of cognitive impairment (CI) on the ADReSS test data. The best scores for each indicator are highlighted in boldface.

| CI Indicator | Detector | Precision | Recall | F1 | Accuracy | Balanced Accuracy |
|---|---|---|---|---|---|---|
| Filler Speech | Keywords | **0.941** | 0.935 | **0.938** | **0.968** | **0.957** |
| | LLM | 0.623 | **0.941** | 0.750 | 0.837 | 0.871 |
| Repetitive Speech | Unigrams | **0.557** | **0.957** | **0.704** | **0.937** | **0.946** |
| | LLM | 0.407 | **0.957** | 0.571 | 0.888 | 0.919 |
| Substitution Errors | MLM | 0.049 | **0.720** | 0.093 | 0.402 | 0.554 |
| | LLM | **0.107** | 0.640 | **0.184** | **0.759** | **0.702** |
| Vague Speech | Keywords | 0.032 | **0.875** | 0.061 | 0.637 | 0.755 |
| | LLM | **0.061** | **0.875** | **0.115** | **0.817** | **0.846** |

###### 4.1.2. *Speech Delays*

The performance of the silence detector for identifying speech delays in the ADReSS test data is presented in Figure 3. The figure shows the trend in precision, recall, F1-score, and accuracy across a range of choices for the minimum silence length hyperparameter while, using the optimal -55 dBFS silence threshold that was determined on the ADReSS training data.

As the minimum silence length increases, the detector becomes more conservative, only flagging long pauses, causing precision to steadily increase while the recall falls. The highest F1-score (35%) was achieved with a minimum silence length of 1,950ms.

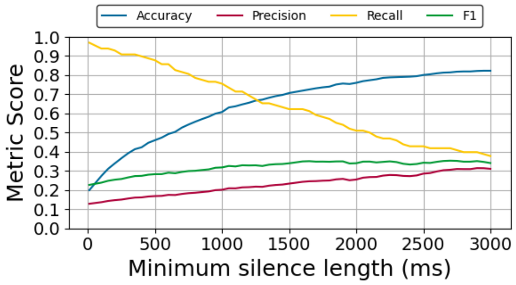


Fig. 3: Performance of silence detector for identifying speech delays in the ADReSS test data.

##### 4.2. *Performance of the Cognitive Impairment Prediction Model*

Figure 4 shows the performance characteristics of the best performing CI prediction model on the ADReSS training and test datasets. The ROC curves and AUC estimates demonstrate the model's strong ability to discriminate between the cognitively impaired and normal groups.

The model achieves a near-perfect AUC of 95% (CI: [0.905, 0.976]) on the training data, and generalized fairly well on the test data (AUC = 80%, CI: [0.682, 0.904]). The model also demonstrates strong F1-scores on both datasets, 87% (CI: [0.792, 0.935]) and 77% (CI: [0.647, 0.867]), on the training and test data, respectively. We anticipated a drop in performance due to a small degree of overfitting to the small training sample size. Nonetheless, the strong train and test performance demonstrates that a model is able to learn meaningful patterns for identifying CI, even from features derived from detectors that often favor high recall over precision.

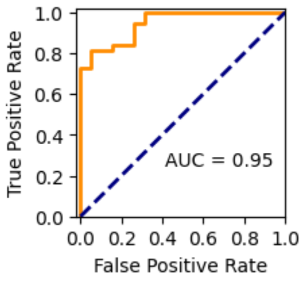


(a) Train

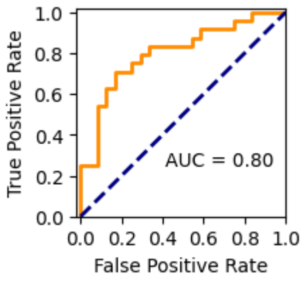


(b) Test

Fig. 4: Performance characteristics of the CI prediction model on the ADReSS datasets.

##### 4.3. *External Validation of WATCH-SS*

We performed an external validation of WATCH-SS on the OBSERVER dataset to assess its viability as an ambient CI screening tool for real-world primary care settings. The CI prediction model's performance on this dataset was lower than on the ADReSS data, achieving an AUC of 63.2% (CI: [0.427, 0.829]) and F1-score of 31.6% (CI: [0.111, 0.526]).

#### Supplementary Material

All appendices can be found at
https://github.com/kbjohnson-penn/WATCH-SS/blob/main/supplementary_material.md.

#### Supplementary Material

##### Appendix A. Keyword Set Optimization for the Filler Speech Detector

To determine the optimal keyword set for the baseline filler keyword detector, we evaluated several combinations of the categories defined in Table A1. We systematically evaluated combinations starting with the highest-performing individual category (Sounds) and progressively adding other categories to assess their complementary value. This approach allows us to balance precision and recall while understanding the marginal contribution of each category. The performance of each configuration on the ADReSS training data is presented in Table A2.

Table A1: Candidate keyword categories for detecting filler speech.

| Category | Description | Keywords |
|---|---|---|
| Sounds | Canonical sounds for filled pauses. | ah, eh, er, hm, huh, mm, uh, um |
| Letters | Single-letter sounds which manifest from stuttering or hesitation. | the alphabet |
| Uncommon Letters | Single-letter sounds excluding those that function as common words. | all letters except a, i, and o |
| Words | Common single-word discourse markers. | like, so, well, actually, basically, literally |
| Non-Words | Phonological fragments or neologisms not in a standard vocabulary, identified as out-of-vocabulary (OOV) tokens. | spaCy Out-of-Vocabulary (OOV) tokens |
| Phrases | Common multi-word discourse markers. | i mean, i guess, you know, you see |

Table A2: Performance of keyword sets for the filler keyword detector on the ADReSS training data. The best scores are highlighted in boldface and the second best scores are underlined.

| Keyword Set | Precision | Recall | F1 | Accuracy | Balanced Accuracy |
|---|---|---|---|---|---|
| Sounds | **0.981** | 0.827 | 0.898 | 0.952 | 0.911 |
| Letters | 0.349 | 0.447 | 0.392 | 0.643 | 0.579 |
| Uncommon Letters | 0.957 | 0.176 | 0.298 | 0.786 | 0.587 |
| Words | 0.341 | 0.122 | 0.179 | 0.713 | 0.520 |
| Non-Words | 0.292 | 0.027 | 0.050 | 0.733 | 0.502 |
| Phrases | 0.276 | 0.031 | 0.056 | 0.730 | 0.501 |
| Sounds + Letters | 0.531 | <u>0.965</u> | 0.685 | 0.772 | 0.835 |
| Sounds + Uncommon Letters | <u>0.975</u> | 0.922 | **0.948** | **0.974** | **0.957** |
| Sounds + Words | 0.772 | 0.835 | 0.802 | 0.894 | 0.875 |
| Sounds + Non-Words | 0.910 | 0.835 | 0.871 | 0.936 | 0.903 |
| Sounds + Phrases | 0.895 | 0.835 | 0.864 | 0.932 | 0.901 |
| Sounds + Uncommon Letters + Words | 0.785 | 0.929 | 0.851 | 0.916 | 0.921 |
| Sounds + Uncommon Letters + Non-Words | 0.911 | 0.925 | <u>0.918</u> | <u>0.958</u> | <u>0.947</u> |
| Sounds + Uncommon Letters + Phrases | 0.843 | 0.929 | 0.884 | 0.937 | 0.935 |
| Sounds + Words + Non-Words | 0.837 | 0.843 | 0.840 | 0.917 | 0.893 |
| Sounds + Non-Words + Phrases | 0.837 | 0.843 | 0.840 | 0.917 | 0.893 |
| Sounds + Uncommon Letters + Non-Words + Words | 0.746 | 0.933 | 0.829 | 0.901 | 0.912 |
| Sounds + Uncommon Letters + Non-Words + Phrases | 0.843 | 0.929 | 0.884 | 0.937 | 0.935 |
| Sounds + Letters + Words + Phrases + Non-Words | 0.482 | **0.973** | 0.644 | 0.724 | 0.805 |
| Sounds + Uncommon Letters + Words + Non-Words + Phrases | 0.703 | 0.937 | 0.803 | 0.882 | 0.900 |

#### Appendix B. Prompt for the Filler Speech LLM-Based Detector

The zero-shot prompt for the LLM-based detector for filler speech is provided in Figure B1. This prompt was selected after an iterative refinement process on the ADReSS training dataset to maximize the F1-score. The performance results are as follows: a F1-score of 74.5%, precision of 62.7%, recall of 91.8%, accuracy 83.9%, and balanced accuracy of 86.4%.

```
# INSTRUCTIONS
You are a neurologist analyzing a patient's speech sample for signs of cognitive impairment.

Your task is to identify the use of filler words in the provided utterance below.

### Definition
Filler words are Sounds (like "uh", "um"), fragments (like "r", "sh"), words (like "well", "like"), or phrases (like "I mean", "you know") used to fill pauses in speech while formulating what to say next. Do not flag word repetition as filler. Do not flag event tags of the format "[<event>]" as filler.

### Output Format
Your output must be a single JSON object with a single key "detections" whose value is an array of JSON objects. Each object in the array represents one detected filler and must have the following keys-value pairs:

  • "type": "filler".
  • "text": The verbatim text from the utterance that was identified as the filler.
  • "span": The character span of the filler in the utterance.

# INPUT
{utterance}
```

Fig. B1: Zero-shot prompt for LLM-based detector for filler words.

#### Appendix C. Hyperparameter Tuning for the Unigram Repetition Detector

We performed a hyperparameter search for the repetitive speech detector based on unigram analysis to determine the optimal configuration that maximizes the F1-score. We evaluated five window sizes (K = 1, 2, 3, 5, and 10) and two comparator functions (an exact match and a lemma-based match, both case-insensitive). The results are shown in Table C1.

Table C1: Hyperparameter search for the word repetition n-gram analysis detector on the ADReSS train data. The best scores are highlighted in boldface and the second best scores are underlined.

| Window Size | Comparator | Precision | Recall | F1 | Accuracy | Balanced Accuracy |
|---|---|---|---|---|---|---|
| 1 | exact | **0.849** | 0.529 | 0.652 | **0.952** | 0.760 |
| 2 | exact | 0.650 | 0.894 | **0.752** | 0.950 | 0.924 |
| 3 | exact | 0.456 | 0.965 | 0.619 | 0.898 | **0.928** |
| 5 | exact | 0.293 | 0.988 | 0.452 | 0.794 | 0.882 |
| 10 | exact | 0.241 | 0.988 | 0.388 | 0.733 | 0.848 |
| 1 | lemma exact | 0.818 | 0.529 | 0.643 | 0.950 | 0.759 |
| 2 | lemma exact | 0.616 | 0.906 | 0.733 | 0.943 | 0.926 |
| 3 | lemma exact | 0.409 | 0.976 | 0.576 | 0.877 | 0.922 |
| 5 | lemma exact | 0.270 | **1.000** | 0.425 | 0.768 | 0.873 |
| 10 | lemma exact | 0.230 | **1.000** | 0.374 | 0.713 | 0.843 |

#### Appendix D. Prompt for the Repetitive Speech LLM-Based Detector

Figure D1 presents the few-shot prompt for the LLM-based detector for word repetitions. We iteratively refined the word repetition definition and rules of exclusion (*i.e.,* what not to detect as repetition) for the prompt in order to maximize the F1-score on the ADReSS traning data. The final prompt performed as follows: 67.8% F1-score, 53.4% precision, 92.9% recall, 92.4% accuracy, and 92.7% balanced accuracy.

```
# INSTRUCTIONS
You are a neurologist analyzing a patient's speech sample for signs of cognitive impairment.

Your task is to identify all clinically significant instances of word repetition in the provided input utterance below.

### Definition
Word repetition is involuntary, immediate verbatim repeat of a word that disrupts the flow of speech and signals a potential struggle with speech production. Do not flag words that repeat for valid grammatical reasons (e.g., "I knew that that was the problem.") or for emphasis (e.g., "very very"). Do not flag word repetitions that are part of a larger phrasal revision (e.g., in "He went to the store [silence] the bank", the repetition of "the" should be ignored).

### Output Format
Your output must be a single JSON object with a single key "detections" whose value is an array of JSON objects. Each object in the array represents one detected repetition and must have the following four keys-value pairs:

  • "type": "repetition".
  • "text": The verbatim text of the repeated word or phrase.
  • "span": The character index for the first occurrence of "text" in the provided input.
  • "span2": The character index for the second occurrence of "text" in the provided input.

### Examples
**Input**: two s uh two cups and a plate are on the um counter there .
**Correct Output**:
{
    "detections": [ {"type": "repetition", "text": "two", "span": [0, 3], "span2": [9, 12]} ]
}

**Input**: and he has a cookie in each hand, handing about to hand one cookie to the little girl who is standing there with her hand reached up for the cookie .
**Incorrect Output**:
{
    "detections": [
        {"type": "repetition", "text": "hand", "span": [28, 32], "span2": [52, 56]},
        {"type": "repetition", "text": "cookie", "span": [14, 20], "span2": [61, 67]}
    ]
}
**Error**: These are not immediate repetitions but rather normal word reuse in different contexts.

**Input**: and her brother's taking cookies out of the cookie jar .
**Incorrect Output**:
{
    "detections": [ {"type": "repetition", "text": "cookie", "span": [37, 43], "span2": [51, 57]} ]
}
**Error**: "cookies" and "cookie" are different word forms and should not be flagged as repetitions.

# INPUT
{utterance}
```

Fig. D1: Few-shot prompt for LLM-based detector for repetitive speech.

#### Appendix E. Percentile Cutoff Optimization for Substitution Errors MLM-Based Detector

Table E1 shows the performance of the MLM-based detector for finding substitution errors using various percentiles for the cutoff threshold for the fusion scores. The results demonstrate the classic precision-recall tradeoff: lower thresholds (90th percentile) maximize recall at the cost of precision, while higher thresholds become too restrictive, missing errors entirely. Substitution errors are scarce in the data so we select the threshold with highest recall, namely the 90th percentile, to maximally detect annotated errors.

Table E1: Evaluation of percentile cutoffs for fusion score for the substitution error MLM-based detector on the ADReSS training data. The best scores are highlighted in boldface and the second best scores are underlined.

| Threshold Percentile | Precision | Recall | F1 | Accuracy | Balanced Accuracy |
|---|---|---|---|---|---|
| 90 | 0.047 | **0.771** | 0.089 | 0.445 | **0.602** |
| 92.5 | 0.049 | 0.657 | **0.092** | 0.542 | 0.597 |
| 95 | **0.051** | 0.486 | **0.092** | 0.662 | 0.577 |
| 98 | 0.030 | 0.114 | 0.047 | 0.838 | 0.489 |
| 99 | 0.029 | 0.057 | 0.039 | **0.900** | 0.494 |

#### Appendix F. Prompt for the Substitution Errors LLM-Based Detector

Figure F1 shows the final zero-shot prompt for the substitution errors LLM-based detector. The final prompt performed as follows on the ADReSS training data: 19.8% F1-score, 11.4% precision, 74.3% recall, 78.7% accuracy, and 76.6% balanced accuracy.

### INSTRUCTIONS
You are a neurologist analyzing a patient's speech sample for signs of cognitive impairment.

Your task is to identify all substitution errors in a patient's speech provided in the input below.

##### Definition
Substitution errors occur when a person involuntarily replaces their intended word with an unintended word while speaking. Focus on detecting the following five substitution errors types:

- Phonemic paraphasias, where sounds within the intended word are added, dropped, substituted, or rearranged (e.g., saying "papple" for "apple").
- Semantic paraphasias, where the intended word is substituted entirely with another real word (e.g., saying "cat" for "dog").
- Neologisms, where the entire intended word is substituted with a non-word (e.g., saying "foundament" for "foundation").
- Morphological errors, where the intended word is used in the incorrect form, such as the wrong number (e.g., saying "child" for "children") or tense (e.g., saying "walked" for "walk").
- Intra-word dysfluencies, where the production of the intended word is disrupted by an inserted sound (e.g., saying "beuhcause" for "because").

Only flag single words that are clinically significant substitution errors and use the surrounding utterances to better understand the context of any given word.

##### Output Format
Your output must be a single JSON object with a single key "detections" whose value is an array of JSON objects. Each object in the array represents one detected substitution error and must have the following key-value pairs:

- "type": "substitution error".
- "text": The verbatim text of the detected substitution error.
- "span": The character span of "text" in the provided input.
- "justification": A brief explanation of why the "text" is a substitution error within its specific context.

### INPUT
{transcript}

Fig. F1: Zero-shot prompt for LLM-based detector for substitution errors.

#### Appendix G. Keyword Set Optimization for the Vague Speech Detector

We considered two categories of vague key words and phrases defined in Table G1, and their combination, to determine which is the optimal keyword set for the baseline vague speech detector. Table G2 shows the performance of the detector across these keyword set combinations.

Table G1: Key words and phrases for vague speech.

| Category | Description | Keywords |
|---|---|---|
| Non-Specific Referents | Words that refer to people, places, or things in vague or general terms without being specific. | anybody, anyone, anything, area, everything, here, it, nothing, one, ones, part, place, people, person, that, there, thing, things, this, someone, somebody, something, stuff, whatever, whichever |
| Hedges | Words and phrases that express uncertainty, approximation, or qualification. | basically, maybe, probably, possibly, potentially, perhaps, somewhat, i guess, i think, kind of, pretty much, sort of, i don't know, more or less |

Table G2: Performance of different keyword set configurations for the vague speech keyword detector on the ADReSS training data. The best scores are highlighted in boldface and the second best scores are underlined.

| Keyword Set | Precision | Recall | F1 | Accuracy | Balanced Accuracy |
|---|---|---|---|---|---|
| Non-specific Referents | **0.031** | **0.917** | **0.060** | <u>0.654</u> | **0.784** |
| Hedges | 0.013 | 0.083 | 0.022 | **0.910** | 0.502 |
| Non-specific Referents + Hedges | <u>0.028</u> | **0.917** | <u>0.054</u> | 0.610 | <u>0.762</u> |

#### Appendix H. Prompt for the Vague Speech LLM-Based Detector

The few-shot prompt for the vague speech LLM-based detector is provided in Figure H1. This vague speech definition, exclusions, and few-shot examples for the prompt were iteratively refined to maximize the F1-score on the ADReSS training data. This prompt achieves 5% precision, 75% recall, 9.4% F1, 82.5% accuracy, and 78.8% balanced accuracy.

```
# INSTRUCTIONS
You are a neurologist analyzing a patient's speech sample for signs of cognitive impairment.

Your task is to identify all instances of vague terms or phrases in a patient's speech provided in the input below.

### Definition
Vague speech occurs when a person struggles to retrieve specific, concrete words and opts to use general placeholders or less specific terms instead. However, do not flag a vague term or phrase if its meaning is either made clear by the context (e.g., it has a specific antecedent) or if it represents a common and appropriate pattern of casual speech that does not indicate word-finding difficulty or uncertainty. Do not flag filler or event and inaudible tokens as vague terms. Do not flag an entire utterance as vague if only a few words or phrases are the source of the vagueness.

### Output Format
Your output must be a single JSON object with a single key "detections" whose value is an array of JSON objects. Each object in the array represents one detected vague speech segment and must have the following key-value pairs:
- "type": "vague".
- "text": The verbatim text of the vague speech detection.
- "utt_num": The utterance number where "text" was detected.
- "span": The character span of "text" in the "utt_num" utterance.

### Examples
**Input**:
5: Provider: okay tell me what else you see .
6: Patient: [silence] some little knots or somethin .
7: Patient: I don't know .
8: Patient: [silence] [inaudible] .
9: Patient: [silence] some kind of a [inaudible] pan or somethin .
10: Patient: [silence] and that girl is there .

**Correct Output**:
{
    "detections": [ {"type": "vague", "text": "or somethin", "utt_num": 6, "span": [32, 42]},
                    {"type": "vague", "text": "or somethin", "utt_num": 9, "span": [41, 51]} ]
}

**Incorrect Output**
{
    "detections": [ {"type": "vague", "text": "[silence]", "utt_num": 8, "span": [0, 9]},
                    {"type": "vague", "text": "that girl", "utt_num": 10, "span": [32, 42]} ]
}
Error: Incorrectly flags (1) "that girl" which was used to identify a person by age and gender rather than express uncertainty about who this person is, and (2) annotation markers ([silence]).

# INPUT
{transcript}
```

Fig. H1: Few-shot prompt for LLM-based detector for vague speech.

#### Appendix I. Hyperparameter Tuning for the Speech Delays Detector

We tune the silence threshold for the speech delays detector to maximize recall the ADReSS training dataset, while fixing the minimum silence length to 10ms. I1Figure I1 shows the results, demonstrating that a threshold of -55dBFS achieves the best recall.

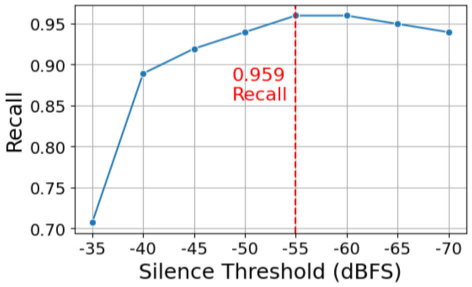


Fig. I1: Performance of the silence detector at identifying speech delays for varying choice of minimum silence threshold on the ADReSS training data.

#### Appendix J. Univariate Analysis of Summary Features

Univariate tests for each summary feature from Section 3.4 was performed on the the ADReSS training dataset to assess the individual utility of each summary feature. These feature values were computed from the outputs of the NLP baseline detectors. For each feature, we tested for statistically significant differences between the cognitively impaired (AD) and cognitively normal (control) groups using both the independent t-test and the Mann-Whitney U test. We also compute the area under the receiver operating curve (AUC) to assess the predictive capability of each feature independently. For the speech delay features, we report results for the three choices of minimum silence length that yielded the most statistically significant Mann-Whitney U test results. The results are shown in Table J1.

Table J1: Univariate analysis of summary metrics for cognitive impairment indicator detectors outputs on the ADReSS training data. Values in the AD Group and Control Group columns are mean (standard deviation). Statistical test results are presented as statistic (p-value).

| Indicator Category | Summary Metric | AD Group | Control Group | T-test | Mann-Whitney U test | AUC |
|---|---|---|---|---|---|---|
| Filler Speech | Filler Rate | 4.49 (5.38) | 3.29 (2.37) | -7.01 (<0.001) | 690.00 (<0.001) | 0.50 |
| | Mean IFD | 3.83 (3.99) | 3.77 (5.81) | -2.50 (0.013) | 1962.50 (0.013) | 0.41 |
| | Standard Deviation IFD | 1.20 (1.82) | 1.25 (1.29) | 0.16 (0.874) | 3490.00 (0.874) | 0.51 |
| Repetitive Speech | Repetition Rate | 2.70 (2.92) | 1.22 (1.27) | -5.26 (<0.001) | 1565.00 (<0.001) | 0.65 |
| | POS Repetition Rate | | | | | |
| | *Adjective* | 0.03 (0.21) | 0.00 (0.00) | 7.86 (<0.001) | 4144.00 (<0.001) | 0.51 |
| | *Adposition* | 0.16 (0.37) | 0.06 (0.26) | 5.56 (<0.001) | 3918.00 (<0.001) | 0.56 |
| | *Adverb* | 0.03 (0.15) | 0.00 (0.00) | 8.10 (<0.001) | 4181.00 (<0.001) | 0.51 |
| | *Auxiliary* | 0.24 (0.48) | 0.25 (0.51) | 3.04 (0.003) | 3507.00 (0.003) | 0.50 |
| | *Coordinating conjunction* | 0.11 (0.30) | 0.08 (0.28) | 5.96 (<0.001) | 3974.00 (<0.001) | 0.53 |
| | *Determiner* | 0.00 (0.00) | 0.00 (0.00) | 8.49 (<0.001) | 4200.00 (<0.001) | 0.50 |
| | *Interjection* | 0.30 (0.92) | 0.10 (0.31) | 3.00 (0.003) | 3769.00 (0.003) | 0.54 |
| | *Noun* | 0.32 (1.25) | 0.01 (0.08) | 2.80 (0.006) | 3993.00 (0.006) | 0.57 |
| | *Numeral* | 0.00 (0.00) | 0.00 (0.00) | 8.49 (<0.001) | 4200.00 (<0.001) | 0.50 |
| | *Particle* | 0.06 (0.23) | 0.04 (0.21) | 6.99 (<0.001) | 4031.00 (<0.001) | 0.51 |
| | *Pronoun* | 1.16 (1.06) | 0.53 (0.63) | -2.86 (0.005) | 2273.00 (0.005) | 0.67 |
| | *Proper noun* | 0.03 (0.17) | 0.00 (0.00) | 8.01 (<0.001) | 4144.00 (<0.001) | 0.51 |
| | *Subordinating conjunction* | 0.00 (0.00) | 0.03 (0.18) | 7.99 (<0.001) | 4144.00 (<0.001) | 0.49 |
| | *Verb* | 0.14 (0.41) | 0.12 (0.35) | 5.03 (<0.001) | 3807.00 (<0.001) | 0.51 |
| | *Other* | 0.10 (0.35) | 0.00 (0.00) | 6.81 (<0.001) | 4050.00 (<0.001) | 0.55 |
| Substitution Errors | Substitution Error Rate | 10.51 (2.62) | 9.53 (3.01) | -28.51 (<0.001) | 0.00 (<0.001) | 0.60 |
| | Mean ISED | 3.38 (1.97) | 3.48 (2.29) | 0.57 (0.571) | 2648.00 (0.571) | 0.50 |
| | Standard Deviation ISED | 1.99 (1.77) | 1.84 (1.91) | -5.21 (<0.001) | 1808.50 (<0.001) | 0.51 |
| Vague Speech | Vague Term Rate | 7.52 (3.76) | 4.62 (2.71) | -13.40 (<0.001) | 224.00 (<0.001) | 0.75 |
| | Vague Utterance Ratio | 39.71 (19.01) | 29.17 (14.52) | -16.67 (<0.001) | 224.00 (<0.001) | 0.67 |
| Speech Delays | Normalized Silence Duration | | | | | |
| | *Min. silence len. = 6,100ms* | 0.11 (0.17) | 0.06 (0.23) | -1.16 (0.249) | 524.5 (0.014) | 0.37 |
| | *Min. silence len. = 6,950ms* | 0.1 (0.16) | 0.05 (0.23) | -0.99 (0.325) | 532.0 (0.014) | 0.38 |
| | *Min. silence len. = 8,800ms* | 0.06 (0.13) | 0.05 (0.23) | -0.43 (0.669) | 558.0 (0.016) | 0.40 |
| | Normalized Silence Count | | | | | |
| | *Min. silence len. = 1,250ms* | 0.00 (0.00) | 0.00 (0.00) | -2.61 (0.011) | 469.0 (0.013) | 0.33 |
| | *Min. silence len. = 6,100ms* | 0.00 (0.00) | 0.00 (0.00) | -1.96 (0.054) | 524.5 (0.014) | 0.37 |
| | *Min. silence len. = 7,050ms* | 0.00 (0.00) | 0.00 (0.00) | -1.65 (0.104) | 533.0 (0.014) | 0.38 |
| | Long-to-Short Silence Ratio | | | | | |
| | *Min. silence len. = 250ms* | 0.53 (0.37) | 0.34 (0.38) | -2.18 (0.032) | 500.0 (0.028) | 0.36 |
| | *Min. silence len. = 400ms* | 0.49 (0.37) | 0.31 (0.37) | -2.08 (0.041) | 506.0 (0.032) | 0.36 |
| | *Min. silence len. = 1,050ms* | 0.36 (0.36) | 0.21 (0.34) | -1.97 (0.053) | 521.0 (0.035) | 0.37 |

#### Appendix K. Performance of Cognitive Impairment Prediction Models

Table K1 provides a comprehensive performance comparison of all seven machine learning models evaluated to predict cognitive impairment (*i.e.*, the "AD" label) on the ADReSS data. For all models, we report the scores for the best-performing hyperparameter configuration determined through manual tuning. The training performance scores are based on the ensembled cross-validation predictions. The LightGBM model performs the best for all metrics in the training dataset, thus we use this as the primary model for WATCH-SS. This model also achieves the most balanced performance on the test data.

Table K1: Evaluation of candidate models for predicting cognitive impairment on the ADReSS datasets. The best value is highlighted in boldface and the second best value is underlined.

| Model | CV | Train | | | | Test | | | |
|---|---|---|---|---|---|---|---|---|---|
| | | Precision | Recall | F1 | Accuracy | Precision | Recall | F1 | Accuracy |
| Logistic Regression | LOO | 0.706 | 0.649 | 0.676 | 0.693 | 0.727 | 0.667 | 0.696 | 0.708 |
| | RSKF | 0.686 | 0.621 | 0.628 | 0.67 | 0.727 | 0.667 | 0.696 | 0.708 |
| Random Forest | LOO | 0.647 | 0.595 | 0.620 | 0.640 | 0.708 | 0.708 | 0.708 | 0.708 |
| | RSKF | 0.737 | 0.664 | 0.678 | 0.713 | 0.667 | 0.667 | 0.667 | 0.667 |
| HGBC | LOO | 0.730 | 0.730 | 0.730 | 0.733 | 0.704 | **0.792** | 0.745 | 0.729 |
| | RSKF | 0.637 | 0.657 | 0.633 | 0.654 | 0.655 | **0.792** | 0.717 | 0.688 |
| LightGBM | LOO | **0.938** | **0.811** | **0.870** | **0.880** | **0.783** | <u>0.750</u> | **0.766** | **0.771** |
| | RSKF | 0.755 | 0.702 | 0.697 | 0.724 | <u>0.750</u> | <u>0.750</u> | <u>0.750</u> | <u>0.750</u> |
| XGBoost | LOO | <u>0.933</u> | <u>0.757</u> | <u>0.836</u> | <u>0.853</u> | 0.704 | **0.792** | 0.745 | 0.729 |
| | RSKF | 0.747 | 0.639 | 0.661 | 0.708 | 0.747 | 0.639 | 0.661 | 0.708 |
| K-Nearest Neighbors | LOO | 0.655 | 0.514 | 0.576 | 0.627 | 0.733 | 0.458 | 0.564 | 0.646 |
| | RSKF | 0.729 | 0.550 | 0.600 | 0.655 | 0.733 | 0.458 | 0.564 | 0.646 |
| Support Vector Machine | LOO | 0.686 | 0.649 | 0.667 | 0.680 | <u>0.750</u> | 0.625 | 0.682 | 0.708 |
| | RSKF | 0.718 | 0.658 | 0.668 | 0.702 | <u>0.750</u> | 0.625 | 0.682 | 0.708 |

#### Appendix L. LightGBM Feature Importance: SHAP Analysis

Figure L1 illustrates the SHAP (SHapley Additive exPlanations) values for our LightGBM model in a beeswarm plot, which provides a comprehensive summary of feature importance across all Leave-One-Out cross-validation folds. The plot reveals that the five most impactful features are related to the vague term rate, followed by the mean inter-filler distance (IFD), repetition rate, the standard deviation of inter-substitution error distance, and the normalized silence count of 1,250 milliseconds. As seen in the plot, high values for vague term rate, repetition rate, and normalized silence 1,250 (red dots on the right) strongly push the model toward a positive CI prediction. Conversely, low values for mean IFD and standard deviation of inter substitution error distance (ISED) tend to increase the model's confidence in a CI prediction. These results confirm the clinical relevance of our chosen indicators, showing that the model's predictions are primarily driven by the same linguistic and acoustic signs that clinicians would look for.

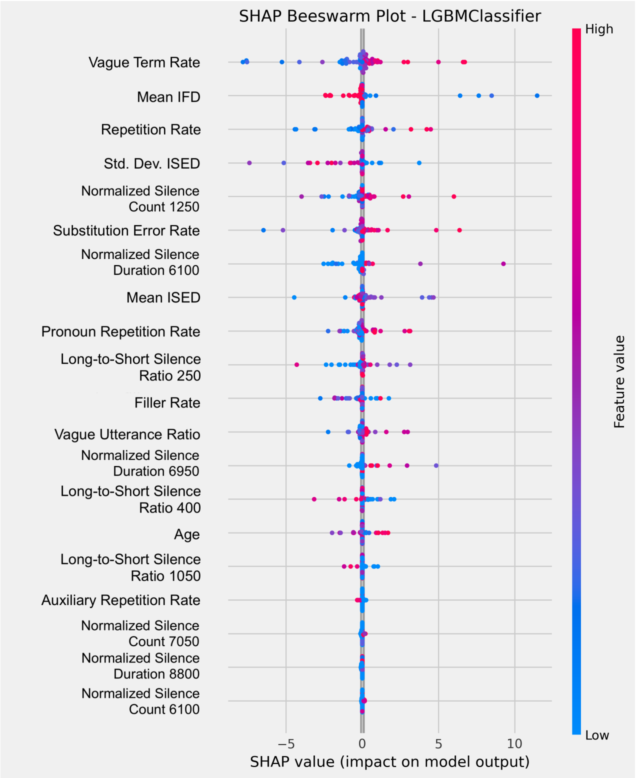


Fig. L1: SHAP beeswarm plot for the LightGBM classifier.
